# Molecular residual disease detection by serial tumour-informed circulating tumour DNA analysis in resectable oesophageal & gastroesophageal junctional adenocarcinoma: a prospective UK multi-centre study

**DOI:** 10.64898/2026.07.06.26357371

**Authors:** Hannah R. Coles, Adam Freeman, Daniel H. Jacobson, Ginny Devonshire, Nicola Grehan, Curtis Millington, Barbara Nützinger, Alice Harvey, John H. Saunders, James Gossage, Rujun Ma, Lucie Mason, Simon L. Parsons, Victoria Askinyte, Spyridoula Massia, Rebecca C. Fitzgerald, Christopher M. Jones

## Abstract

**Background:** The value and optimal timing of circulating tumour DNA (ctDNA) analysis in locally advanced oesophageal adenocarcinoma (OAC) is uncertain. We hypothesised that perioperative detection would be prognostic for event-free (EFS) and overall (OS) survival, and that 3-6 months post-operative detection would identify recurrence earlier than standard-of-care surveillance.

**Methods:** In this prospective exploratory multi-centre cohort study, tumour-informed ctDNA assays were designed for 49 patients using whole-exome sequencing. Bloods were collected for ctDNA detection up to 8 days prior to surgery, and at 3-6 weeks and 3-6 months post-surgery, then correlated with clinicopathological characteristics, EFS and OS.

**Results:** Pre-and early post-surgery ctDNA positivity were associated with worse EFS (HRs 7.97 (95% confidence interval, CI 2.64-24.04), p<0.0001; 8.18 (95%CI 3.23-20.69), p<0.0001) and OS (HRs 7.82 (95%CI 2.22-27.54), p=0.00018; 13.69 (95%CI 4.52-41.49), p<0.0001). In pre-surgery positive patients, post-surgery ctDNA clearance was associated with improved EFS and OS, and was prognostic for OS in patients whose tumours had a poor histopathological response (tumour regression grade, TRG, 4-5). 3-6 month ctDNA-positivity preceded standard-of-care recurrence detection by median 53.5 (interquartile range 39.3-200.0) days.

**Conclusions:** Perioperative ctDNA positivity was associated with worse EFS and OS in OAC, and persistent negativity identified patients with the best outcomes. Post-surgery ctDNA status carried prognostic information not captured by pathological stage or TRG. ctDNA testing at 3-6 months identified patients at high risk of recurrence or death.

## BACKGROUND

Oesophageal adenocarcinoma (OAC) is a major cause of cancer-related morbidity and mortality. Global incidence averages 0.9 cases per 100,000 persons, though rates reach as high as 3.4 cases per 100,000 persons in North America, Europe and Australasia^1^. Approximately 30-40% of patients present with potentially curable locally advanced disease^2^. However, only around half of these patients will survive to five years despite intensive, morbid multi-modality treatment^3^. There is at present limited ability to personalise the management of these patients based on their likely prognosis. This results in under- and over-treatment, with deleterious consequences for cancer survival and health-related quality of life.

Surgical resection is the cornerstone of curative treatment for locally advanced OAC but perioperative therapy improves outcomes^4^. As established by the ESOPEC trial, standard perioperative therapy for most patients is four pre- and four post-operative cycles of 5-fluorouracil and leucovorin, oxaliplatin and docetaxel (FLOT)^3^. However, the 2025 publication of initial results from the MATTERHORN trial provides evidence for an increase in the proportion of patients with pathological complete response (pCR; 19.2% vs. 7.2%), as well as in two-year overall survival (75.8% vs. 70.4%), following perioperative programmed death ligand 1 inhibition using durvalumab^5^. The importance of the adjuvant component of FLOT is contentious. It is not clear that all patients who currently receive post-operative FLOT benefit from it and this treatment adds toxicity ^6,7^. Reflecting this, in the ESOPEC and MATTERHORN trials, 64.3-73.0% of patients started adjuvant FLOT but this was completed as planned in 48.3-53.4%^3,5^.

There is a corresponding need to identify those most likely to derive benefit from adjuvant therapy. Pathological indices have been studied in this context, with response to neoadjuvant therapy generally categorised by tumour regression grade (TRG), such as through the Mandard criteria^8^. This summarises primary tumour cancer cell content in the context of the degree of regression and fibrosis. There is a higher risk of recurrence in patients with residual primary tumour or nodal disease as well as for those with lymphovascular invasion and minimal downstaging in response to neoadjuvant therapy^9–12^. However, these variables are imperfect and around 5-10% of the overall 10-20% of patients who have no pathologic residual disease in the primary tumour or lymph nodes develop recurrence within two years^3,7^. There may also be a benefit to the early detection of recurrent disease, yet current surveillance protocols generally rely on computed tomography (CT), which is poorly sensitive and can be applied only infrequently^13^.

The detection of circulating tumour DNA (ctDNA) has shown promise as a means by which to detect post-operative molecular residual disease (MRD) across multiple cancers^14^. This includes emerging evidence for the use of ctDNA assays in oesophagogastric adenocarcinomas^15–18^. However, previous analyses have predominantly focussed on cohorts of patients with gastric cancer and are not readily translatable to the clinic due to a reliance on broad sampling points and heterogenous populations of patients managed with diverse therapies.

Here, we present the prognostic value of serial pre- and post-operative tumour-informed ctDNA measurements in an exploratory prospective cohort of patients with oesophageal and gastroesophageal junctional adenocarcinoma.

## METHODS

### Study approach

This prospective cohort study enrolled patients from four English National Health Service centres **(Supplementary Table 1)** between February 2023 and June 2024. Patients aged 16 years or over with a diagnosis of resectable oesophageal or gastroesophageal junctional adenocarcinoma were eligible for inclusion. Potential participants were approached prior to surgical resection and were required to be able to provide written informed consent. Exclusion criteria included an additional cancer diagnosis, human immunodeficiency virus or hepatitis B virus positivity, and pregnancy. Patients with a comorbidity that in the treating clinician’s view would increase the risk from regular blood sampling (e.g. bleeding disorders, severe chronic obstructive pulmonary disease, severe anaemia) were also excluded. Patients must not have had a blood transfusion in the two weeks prior to any blood collection for ctDNA analysis.

Sampling timepoints for ctDNA analysis were pre-surgery (at any point from eight days prior to surgery up to and including the day of surgery, prior to knife to skin), early post-surgery (3-6 weeks following resection) and 3-6 months post-resection. Peripheral blood was collected at each of these timepoints using two 10ml Cell-Free DNA blood collection tubes (Streck, Nebraska, USA). An additional 6ml ethylene diamine tetra acetic acid (EDTA) tube was collected prior to surgery for germline sequencing. Formalin-fixed, paraffin embedded (FFPE) tumour resection specimens were assessed by an expert pathologist at each local site to select representative blocks meeting the criteria of greater than 20% tumour cellularity and at least 25mm^2^ surface area. A single haematoxylin and eosin-stained slide and eight 10µm thick unstained slides were prepared by Addenbrooke’s Hospital Human Research Tissue Bank (Cambridge University Hospitals NHS Foundation Trust, Cambridge, UK) for whole exome sequencing.

Clinicians and patients were blinded to ctDNA results. Patient demographic, tumour, treatment and outcomes information were prospectively collated by each site and shared centrally in linked-anonymised form for analysis. This included local evaluation of TRG by Mandard criteria with no central pathology review.

### Tumour-informed ctDNA analysis

Natera, Inc. (Texas, USA) performed the Signatera™ multiplex polymerase chain reaction-next generation sequencing ctDNA assay. Briefly, genomic DNA obtained from archived FFPE tumour tissue and whole blood from each patient were subjected to whole exome sequencing (WES) to select 16 high-quality somatic single nucleotide variants (SNVs) to design bespoke ctDNA assays. Extracted cell-free DNA (cfDNA) from longitudinal blood samples was processed for ctDNA analysis and cfDNA libraries amplified by mPCR using primers designed to the 16 SNVs and subjected to NGS. Plasma samples were categorized as positive if at least two tumour-specific SNVs out of 16 were detected. ctDNA concentration was reported as mean tumour molecules per ml (MTM/ml) of plasma^19^.

### Statistical analyses & data representation

The primary outcome was the association between ctDNA positivity and event free survival (EFS). This was defined as the time from surgery to clinical or radiographic recurrence, confirmed by assessment by an expert multidisciplinary team (MDT), or death. Secondary outcome measures included overall survival (OS), defined as the interval between the date of surgery and either death from any cause or last known follow-up, and Mandard TRG. For inclusion in analyses patients were required to have completed pre-surgery and early post-surgery sampling. This study is pragmatic and the sample size is not based on an a priori power calculation.

Patient characteristics were summarised using descriptive statistics. A summary of the mutations identified through tumour whole exome sequencing was presented in an oncoplot using the ComplexHeatmap R package. Comparisons of ctDNA values across clinical stages were conducted using two-sided Wilcoxon rank sum tests. All survival analyses, including Kaplan-Meier and Cox proportional hazard analyses, were conducted using the survival and survminer R packages. Two sample analyses were performed using Wilcoxon rank sum tests and two-tailed Fisher’s exact test used to compare predictive values. Correlation analyses were performed using Pearson’s correlation coefficients. All analyses were conducted using R (version 4.5.1) and all plots were generated using ggplot2 (version 4.0.2) and ggplot2 extension packages. Hazard ratios reported in the Kaplan-Meier analysis were calculated using unadjusted Cox proportional hazards models, and reported p-values were calculated using a log-rank test. Multivariate Cox proportional hazards modelling included prespecified variables (sex, age, tumour (T-) and nodal (N-) stage according to the 8^th^ AJCC UICC TNM classification system, TRG and, where relevant, receipt of adjuvant chemotherapy) to determine the independent effects of data-driven variables (pre-surgical ctDNA, post-surgery ctDNA, and combined perioperative ctDNA status). Patients with missing data were excluded from relevant multivariate analyses. Statistical significance was established as p<0.05 unless otherwise indicated. False discovery rate (FDR) adjustment for multiple testing correction was conducted across subgroup analysis for event-free and overall survival analysis, and analysis of pre- and post-surgery ctDNA and combined perioperative ctDNA status, separately. Code to reproduce the analysis is available on Github: https://github.com/fitzgerald-lab/Periop_ctDNA.

### Ethics & governance

This study was prospectively approved by the Cambridge South Research Ethics Committee (REC 10/H0305/1). All participants provided written informed consent prior to study enrolment.

## RESULTS

### Study cohort

As outlined in **Figure 1** and summarised in **Supplementary Table 2**, 89 patients consented to study participation. Of these, ctDNA results could not be generated from primary tumour resection specimens for 22.5% (n=20/89) because of insufficient tumour tissue for whole exome sequencing in 10.1% (n=9/89) of the consented cohort and insufficient tumour cellularity due to pCR for 11.2% (n=10/89), as well as failed germline extraction in one patient. For one included patient, a prior endoscopic mucosal resection sample was used for WES due to insufficient tumour tissue in the resection specimen. Diagnostic biopsy specimens did not otherwise meet size criteria. Lymph node tissue was examined for two of the five patients who had insufficient resection tissue and who had positive nodes at resection but neither sample had sufficient tumour content. An additional 10.1% (n=9/89) of the consented cohort were excluded due to a failure to proceed to curative resection, an inability to complete perioperative blood sampling due to surgical complications, or a change in diagnosis based on analysis of the resection specimen. The remaining exclusions (n=11/89; 12.4%) related to withdrawal of consent or logistical issues relating to sample collection and processing. A summary of the clinicopathological characteristics and treatment outcomes for the patients for whom a ctDNA panel could not be generated are provided in **Supplementary Table 3**.

**Figure 1:**
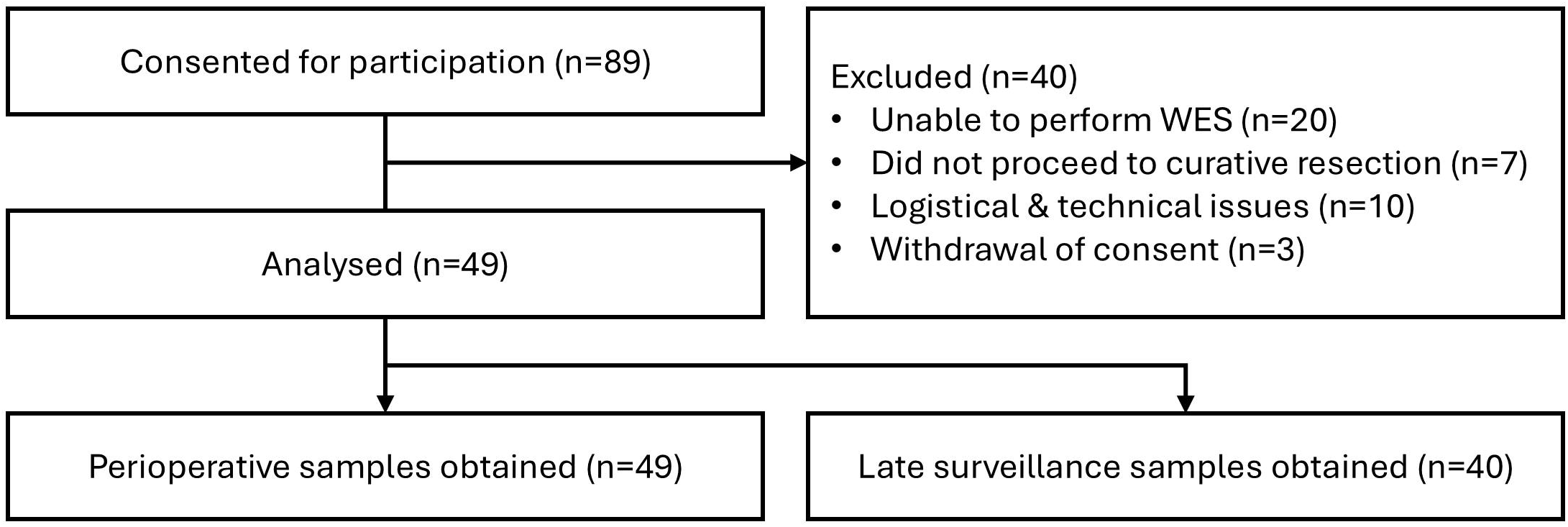
**Consort diagram illustrating identification of the evaluable cohort. WES: whole exome sequencing.**

Following these exclusions, a cohort of 49 evaluable patients with pre-surgical and early post-surgical ctDNA results was established. ctDNA measurements 3-6 months after surgery were available for 81.6% (n=40/49) of the cohort. These patients were mostly male (85.7%; n=42/49) with a median age of 66 (interquartile range, IQR 57-72) years **(Table 1)**. Tumour genomic characteristics were similar to those expected from the general OAC population, with 67% (n=33/49) of patients harbouring a TP53 mutation **(Supplementary Figure 1)**. All patients had microsatellite stable tumours. A summary of neoadjuvant and adjuvant treatments is provided in Supplementary Table 4. Three patients who had cT1b-cT2 cN0 disease did not receive preoperative treatment. Most participants treated with neoadjuvant FLOT had a tumour regression grade of 3 (n=20/46; 43.5%) or 4 (n=21/46; 45.7%) **(Supplementary Table 4)**. Adjuvant FLOT was delivered to 77.6% (n=38/49) of patients, although just over two thirds (n=26/38; 68.4%) received all four planned cycles due to treatment-related adverse events in the other one third.

**Table 1:** A summary of baseline patient and tumour characteristics. *Unknown in one patient. ** Most severe classification shown. Differentiation status unknown in two patients. One patient had M1 disease due to out-of-field nodal involvement. ECOG: Eastern Cooperative Oncology Group.

|  | n=49 | % |
|---|---|---|
| **Patient characteristics** |  |  |
| Age |  |  |
| 40-49 | 5 | 10.2 |
| 50-59 | 10 | 20.4 |
| 60-69 | 17 | 34.7 |
| 70-79 | 16 | 32.7 |
| 80-89 | 1 | 2.0 |
| Sex |  |  |
| Male | 42 | 85.7 |
| Female | 7 | 14.3 |
| ECOG Performance status* |  |  |
| 0 | 32 | 65.3 |
| 1 | 16 | 32.7 |
| **Tumour characteristics** |  |  |
| Tumour location |  |  |
| Lower oesophageal | 9 | 18.4 |
| GOJ |  |  |
| Unclassified | 6 | 12.2 |
| Siewert I | 10 | 20.4 |
| Siewert II | 20 | 40.8 |
| Siewert III | 4 | 8.2 |
| Tumour differentiation** |  |  |
| Moderate | 19 | 38.8 |
| Poor | 28 | 57.1 |
| Clinical T-stage |  |  |
| 1 | 3 | 6.1 |
| 2 | 12 | 24.5 |
| 3 | 28 | 57.1 |
| 4 | 6 | 12.3 |
| Clinical N-stage |  |  |
| 0 | 22 | 44.9 |
| 1 | 13 | 26.5 |
| 2 | 14 | 28.6 |

### ctDNA detection

A total of 138 plasma samples were available for analysis. A summary of ctDNA collection points and their relationship to treatment outcomes is provided in **Figure 2**. Where a panel was generated, results were received a median of 11.4 (IQR 5.3-13.8) days after sample submission to the commercial ctDNA provider. Pre-operative samples were taken on the day of surgery for 91.8% (n=45/49) and between 1-8 days prior for the remaining 8.2%. Half (n=24/49) of the pre-surgical samples had detectable ctDNA, which was identified in 50% (n=23/46) of patients who received neoadjuvant therapy and 33% (n=1/3) of those who did not. Median pre-surgical ctDNA concentration was significantly higher with nodal involvement (p<0.001, q=0.002) and increased with increasing TRG (p<0.001, q<0.001; **Figure 3A**) but there was no association with sex, differentiation status, or anatomical location **(Supplementary Table 5)**. By contrast, there were no significant associations between early post-operative ctDNA positivity and pre-surgical clinical or tumour characteristics, ypT- and ypN-stage, or TRG **(Figure 3B, Supplementary Table 5)**. Early post-surgical and 3-6 month post-surgical ctDNA measurements were taken at a median of 31 (IQR 25-41) and 140 (IQR 113-172) days post-surgery, respectively. There was no association between post-operative clinicopathological characteristics and ctDNA concentration **(Supplementary Table 5)**.

**Figure 2:**
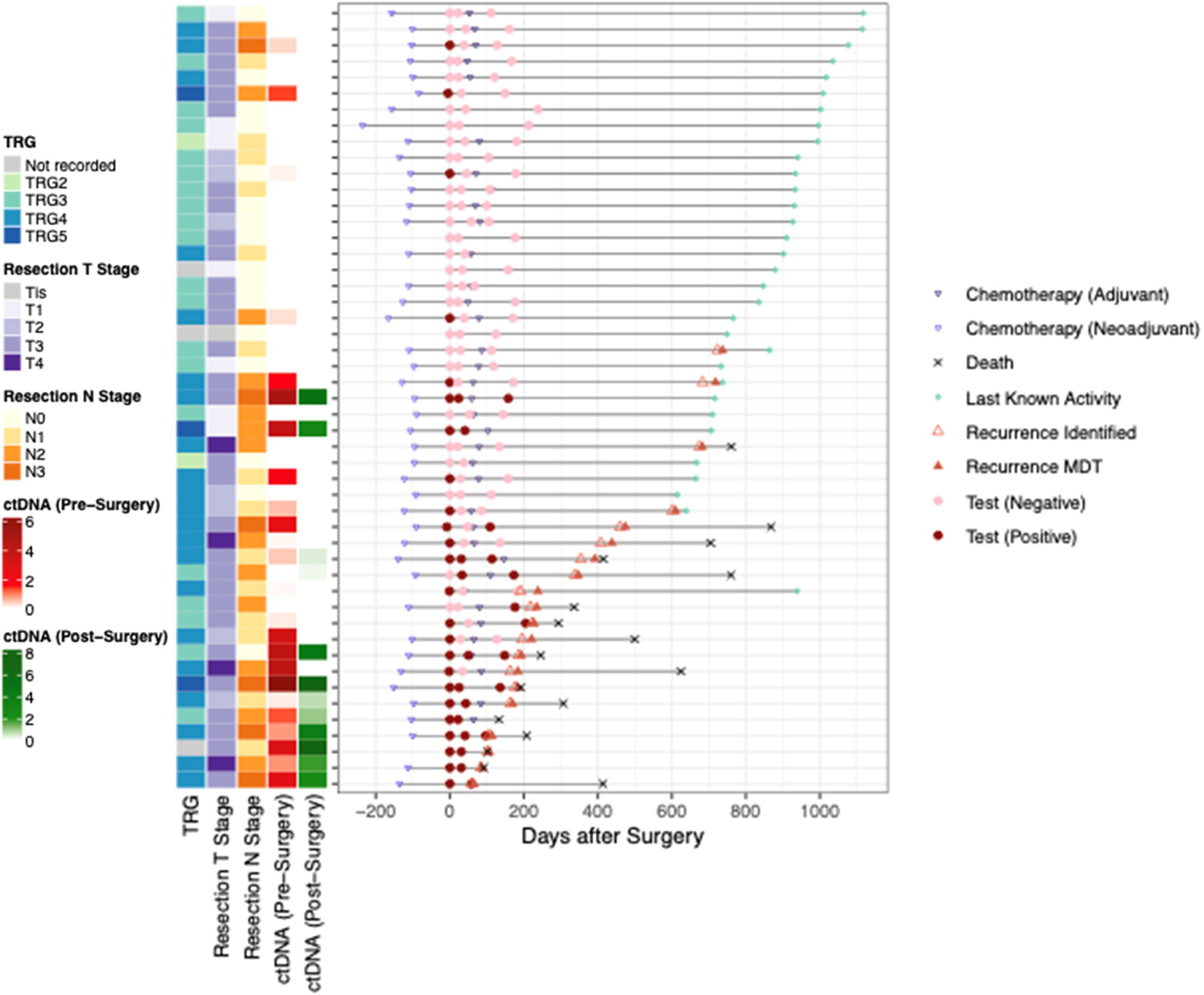
Swimmer plot illustrating the relationship between ctDNA assessment, treatment and outcomes. Patients are ordered by event free survival following surgery. ‘Recurrence identified’ refers to the identification of recurrence via an imaging modality. ‘Recurrence MDT’ refers to formal confirmation of recurrence at a tumour site specific MDT meeting. MDT, multidisciplinary team; TRG, tumour regression grade. Values for ctDNA correspond to log2-normalised mean tumour molecules per ml, with post-surgery value referring to the first test post-surgery.

**Figure 3:**
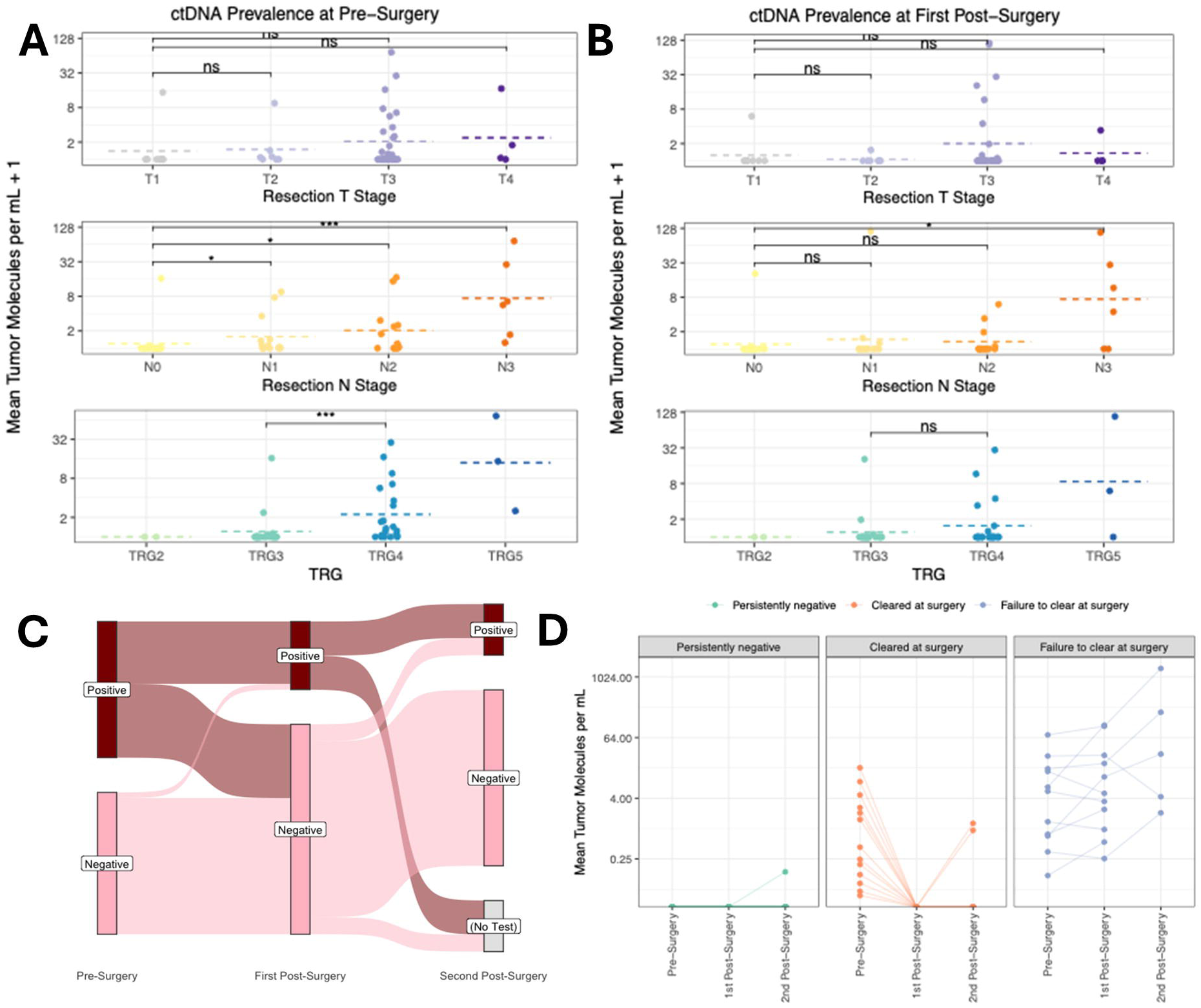
ctDNA positivity by treatment response and over time. (A, **B)** Pre-(A) and post-(B) surgery ctDNA concentration by key pathological variables. P-values were calculated using a two-sided Wilcoxon rank-sum test with Benjamini-Hochberg correction for multiple testing. **(C)** Alluvial plot demonstrating per patient trends in ctDNA positivity at each sampling point. **(D)** Patterns of ctDNA positivity across the sampling timepoints, grouped by ctDNA positivity in perioperative measurements. N-stage: nodal stage; T-stage: tumour stage; TRG: tumour regression grade.

ctDNA dynamics are summarised in Figure 3C. The overall test positivity rate decreased from 49.0% (n=24/49) to 24.5% (n=12/49) at the early post-surgical timepoint and remained similar (22.5%, n=9/40) at the 3-6 month post-surgery timepoint. The per patient trends underlying these proportions are shown in Supplementary Figure 2A-D. As summarised in Figure 3D, these were grouped into three comparable patterns based on pre-surgical and early post-surgical ctDNA measurements: persistently negative (49%; n=24/49), cleared at surgery (27%; n=13/49), and failure to clear at surgery (22%; n=11/49). One patient (2%) became newly positive after surgery, and so was excluded from downstream analyses relating to these perioperative categories. Notably, two (15.4%) of the 13 patients who cleared ctDNA at surgery subsequently returned positive results at the 3-6 month post-surgery timepoint (Supplementary Figure 2B). There was no significant difference in pre-surgery ctDNA concentration between patients who did or did not clear ctDNA at surgery, though the latter was numerically higher (median ctDNA concentrations of 5.52 vs 0.428 MTM/ml; p=0.082; Supplementary Figure 2E). No patients who were ctDNA-positive at the early post-surgery timepoint cleared ctDNA at the 3-6 month post-surgery timepoint.

### Association between perioperative ctDNA detection and outcomes

The median follow-up time was 25.0 (IQR 20.2-30.7) months. During this period, 40.8% (n=20/49) of the cohort developed a recurrence; 25% (n=5/20) of which were locoregional and 75% (n=15/20) of which were metastatic. Overall, 34.7% (n=17/49) of the cohort died, all but one of whom had a known recurrence at the time of death, resulting in an overall event rate of 42.9% (n=21/49). To determine the prognostic value of perioperative ctDNA testing, we evaluated the impact of ctDNA positivity at each time point on EFS and OS. The event rate was 70.8% (n=17/24) with pre-surgery ctDNA positivity versus 16% (n=4/25) with pre-surgery ctDNA negativity (p=0.0001; Fisher’s exact test); correlating with a marked reduction in EFS (unadjusted HR 7.97 (95%CI 2.64-24.04), p<0.0001; **Figure 4A**) and OS (unadjusted HR 7.82 (95%CI 2.22-27.54), p=0.00018; **Figure 4B**). For ctDNA positive patients at the early post-surgery assessment, the event rate was greater still, at 83.3% (n=10/12), compared to 29.7% (n=11/37) for those who were ctDNA negative post-surgery (p=0.002, Fisher’s exact test) (Supplementary Figure 3A). EFS (unadjusted HR 8.18 (95%CI 3.23-20.69), p<0.0001; Figure 4C) and OS (unadjusted HR 13.69 (95%CI 4.52-41.49); p<0.0001; Figure 4D) again differed markedly by positivity at this time point. In those positive for ctDNA, there was no significant correlation between pre-(r=-0.21, p=0.42) or early post-(r=-0.54, p=0.1) surgery ctDNA concentration and time to event **(Supplementary Figure 3C)**.

**Figure 4:**
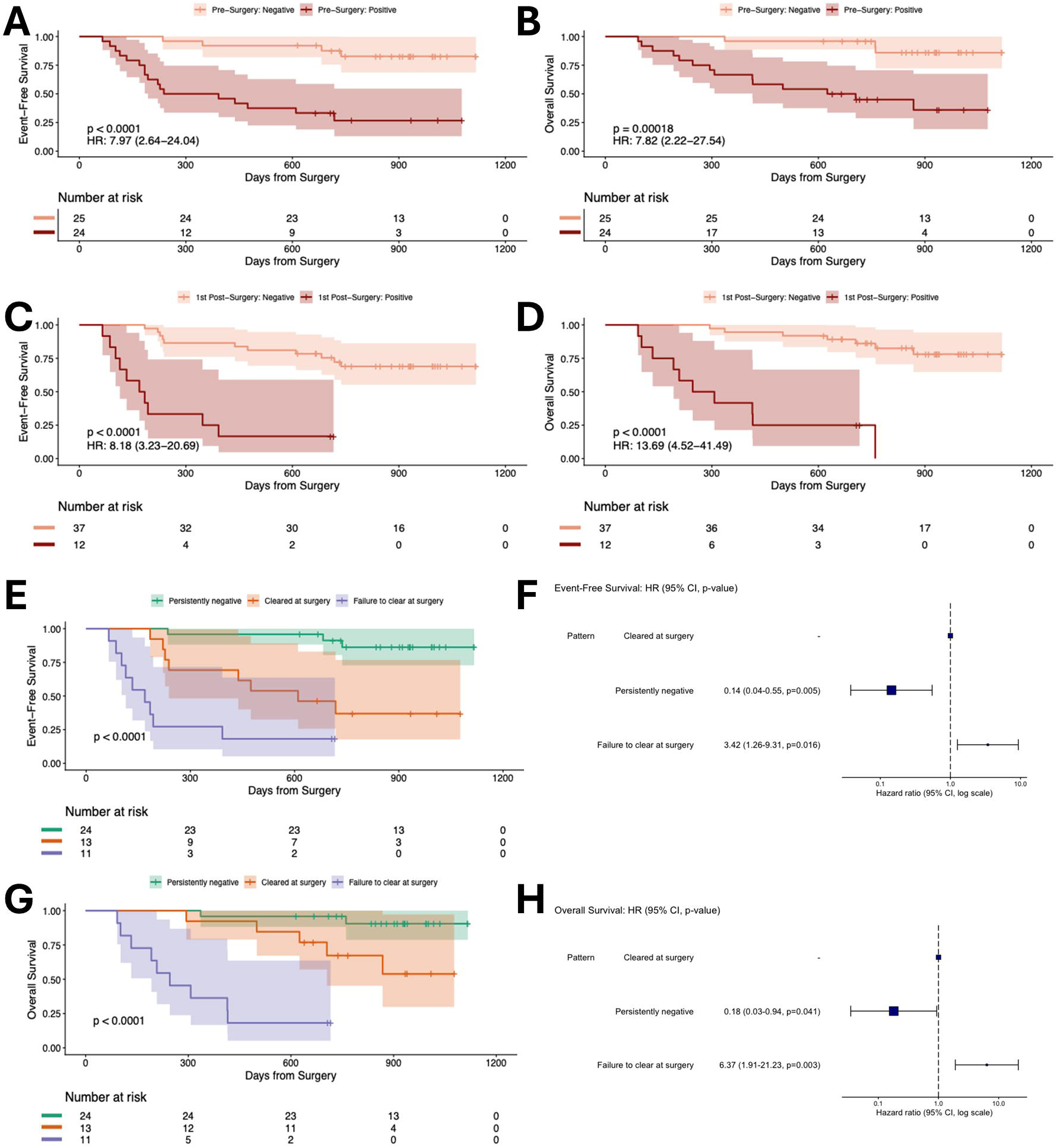
Event-free and overall survival outcomes by sampling timepoint. **(A)** Event-free and **(B)** overall survival by pre-surgical ctDNA status. **(C)** Event-free and **(D)** overall survival by early post-surgery ctDNA status. **(E,F)** Event-free and **(G,H)** overall survival by perioperative (pre-surgery and early post-surgery) ctDNA pattern. Data are not shown for a single patient with newly acquired post-operative ctDNA positivity. P-values were calculated using the log-rank test and underwent FDR-correction for multiple testing across event-free and overall survival analyses separately.

We next evaluated whether the pattern of pre- and post-surgery ctDNA positivity is informative of event risk. Overall, 12.5% (3/24) of patients with persistently negative ctDNA recurred (two with metastatic disease and one with locoregional disease), compared with 61.5% (n=8/13) who cleared ctDNA after surgery (seven with metastatic disease, one with locoregional disease) and 72.7% (n=8/11) who were ctDNA positive after surgery (six with metastatic disease, two with locoregional disease) **(Supplementary Figure 3D)**. One patient who was negative for ctDNA prior to surgery but positive after surgery developed a locoregional recurrence at 11.4 months. Of the three patients with persistently negative perioperative ctDNA who recurred, one had a TRG3 response and ypT3 ypN2 disease after neoadjuvant treatment, and notably developed ctDNA positivity 3-6 months after surgery with recurrence identified at seven months. The other two patients had ypT4 ypN2 disease with a TRG4 response to neoadjuvant therapy, developing a recurrence at 29 months, and ypT3 ypN1 disease with a TRG3 response, recurring at 20 months.

As summarised in **Figures 4E-H**, EFS and OS were significantly lower for patients who did not clear ctDNA after surgery (unadjusted HR 3.42, 95%CI 1.26-9.31, p=0.016 for EFS; unadjusted HR 6.37, 95%CI 1.91-21.23, p=0.003 for OS). EFS and OS were also significantly better for patients who were persistently ctDNA-negative compared to those who converted from ctDNA-positive to ctDNA-negative after surgery (unadjusted HR 0.14, 95%CI 0.04-0.55; p=0.005 for EFS; unadjusted HR 0.18, 95%CI 0.03-0.94, p=0.041 for OS).

In exploratory multivariate analyses, pre-surgery, early post-surgery and combined perioperative ctDNA measurements were independently associated with EFS and OS **(Supplementary Figure 4)**. As shown for combined perioperative measurements in **Figure 5** and individual pre- or early post-surgery measurements in **Supplementary Figure 5**, ctDNA positivity added to the prognostic value of pathological indices. For patients with ypT1/2 tumours, pre-surgical ctDNA positivity associated with event-rate and mortality, regardless of whether ctDNA was cleared at surgery, with no recurrences in patients who had ypT1/2 disease and persistently negative perioperative ctDNA **(Figure 5A, B; Supplementary Figure 5A-D)**. For later stage ypT3/4 tumours, outcomes were again poorest for those with positive pre-surgery ctDNA **(Figure 5A, B; Supplementary Figure 5A-D)**. Compared with those that cleared at surgery, a failure to clear ctDNA at surgery was significantly associated with EFS (HR 4.64, p=0.008) and OS (HR 8.15, p=0.002) **(Supplementary Table 6)**. There were nevertheless still episodes of recurrence in three patients with ypT3/4 disease with persistently negative ctDNA at both pre- and post-surgery timepoints **(Figure 5A)**. These patients all had nodal disease at presentation and at surgical resection, and were found to have a TRG of 3 (n=2) or a TRG of 4 (n=1).

**Figure 5:**
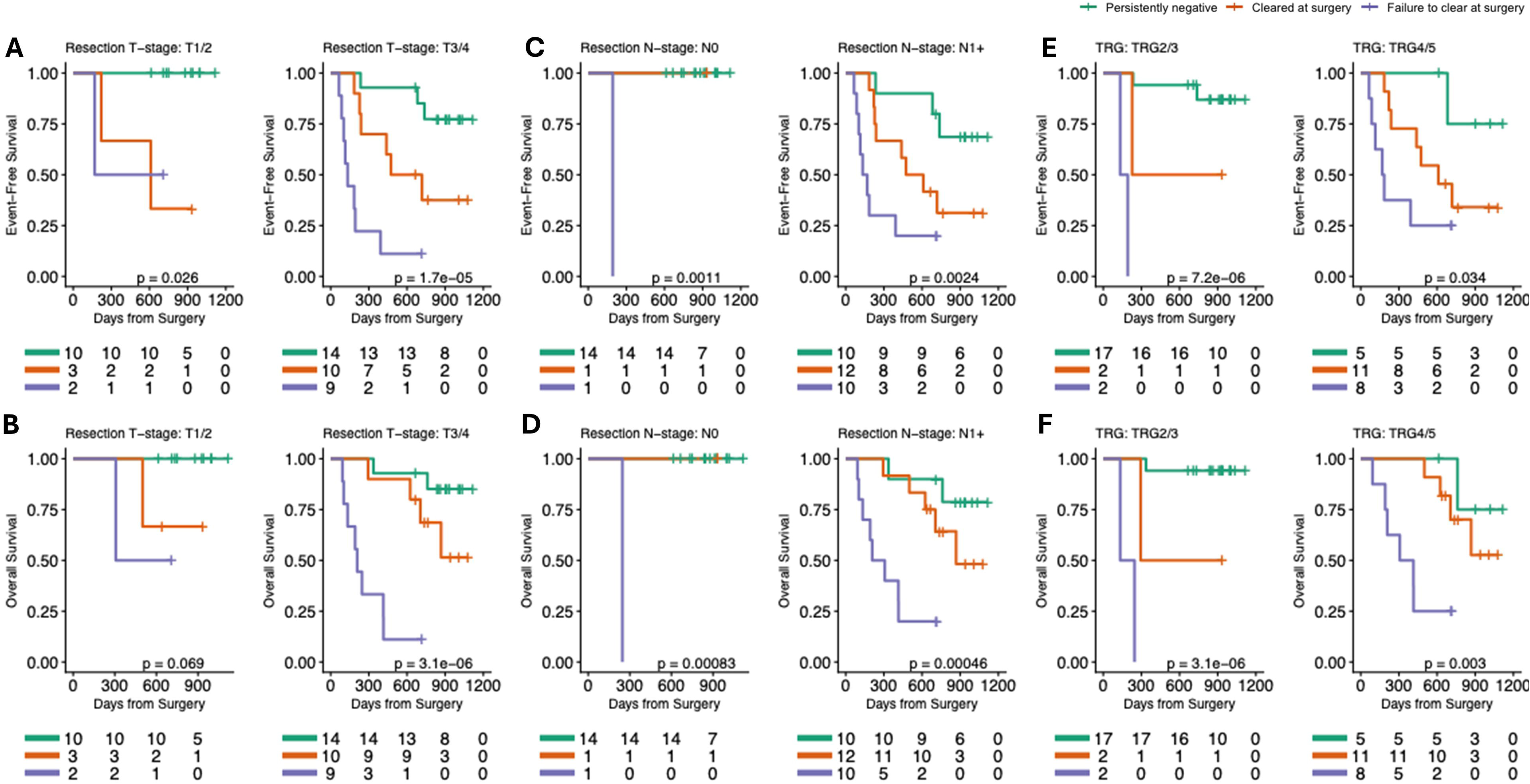
**Event-free and overall survival outcomes by perioperative ctDNA status and pathological indices**. **(A)** Event-free and **(B)** overall survival by perioperative ctDNA pattern and T-stage. **(C)** Event-free and **(D)** overall survival by perioperative ctDNA pattern and N-stage. **(E)** Event-free and **(F)** overall survival by perioperative ctDNA pattern and TRG.

A similar gradient existed for both EFS and OS for patients with nodal involvement at resection in further exploratory multivariate analyses **(Figure 5C, D; Supplementary Figure 5E-H)**. However, there was no recurrence or death in patients with ypN0 disease and negative post-operative ctDNA **(Supplementary Figure 5G, H)**. One patient with ypN0 disease had failure to clear ctDNA after surgery and was identified to have a recurrence, then died, at 6.3 and 8 months post-surgery, respectively. A failure to clear ctDNA at surgery was again significantly associated with event free (unadjusted HR 2.91, p=0.041) and overall (unadjusted HR 5.36, p=0.007) survival in the ypN1+ group **(Supplementary Table 6)**. Pre- and early post-surgery ctDNA assessment also appear to provide additional information over TRG alone, albeit again caveated by small sample sizes and low event rates. Patients who were persistently negative for ctDNA at both perioperative timepoints again had the best EFS and OS, regardless of TRG **(Figure 5E, F; Supplementary Figure 5I-L)**. A failure to clear ctDNA at surgery conferred a greater risk of death in those with TRG4/5 disease (unadjusted HR 6.79, p=0.010; **Supplementary Table 6)**.

Reflecting these trends but caveated by small sample sizes and treatment selection bias, adjuvant chemotherapy appears associated with superior EFS (unadjusted HR 0.16, 95%CI 0.04-0.76, p=0.0098) and OS (unadjusted HR 0.25, 95%CI 0.06-1.03, p=0.039) in patients with a positive early post-surgery ctDNA result **(Supplementary Figure 6A)**. In contrast, survival was numerically better for patients who were ctDNA negative at the early post-operative timepoint and who did not receive adjuvant chemotherapy **(Supplementary Figure 6B)**. These results are confounded by a higher proportion of higher ypT- and ypN-stages in those not receiving adjuvant chemotherapy versus those who did in patients with ctDNA positivity in the post-operative period **(Supplementary Figure 6C)**, with similarly higher proportions of ypT- and ypN-stages in those receiving adjuvant chemotherapy who were ctDNA negative in the postoperative period **(Supplementary Figure 6D)**. These data are also biased by the 11.2% of the consented cohort for whom a panel could not be designed due to pCR.

### Post-operative ctDNA performance

The 3-6 month post-surgery timepoint coincides with usual initiation of surveillance. ctDNA positivity at this timepoint was associated with an 89% (8/9) risk of recurrent disease, versus a 22.6% (n=7/31) risk in those who were ctDNA negative (p=0.0005). In terms of recurrences, a positive ctDNA result at 3-6 month post-surgery occurred at a median of 53.5 (IQR 39.25-200, range 17-365) days before a recurrence was recorded at MDT. Of the eight patients with a positive ctDNA test at this timepoint, 87.5% (n=7/8) had previously had a positive ctDNA test; with 75% (n=6/8) positive pre-surgery and 62.5% (n=5/8) positive early post-surgery. One patient tested negative prior to surgery but positive after surgery.

In contrast, 71.4% (n=5/7) of patients with negative ctDNA results at 3-6 months but who subsequently recurred had a positive test pre-surgery. None were positive early post-surgery. For these patients, recurrence occurred a median of 525 (IQR 198-547, range 55-624) days after the 3-6 month ctDNA test.

## DISCUSSION

In this prospective multi-centre cohort study, we show that perioperative tumour-informed ctDNA testing is feasible in OAC, and provide data to suggest that serial perioperative ctDNA measurement confers additional prognostic value to either pre- or post-surgery ctDNA assessment alone. We also demonstrate that later assessment of ctDNA identifies recurrence significantly earlier than standard surveillance pathways.

Pre-surgery ctDNA concentration associates with ypN-staging and TRG, with a similar albeit non-significant association seen with ypT-stage. Patients who are ctDNA positive pre-surgery have worse EFS and OS than those who are ctDNA-negative. ctDNA clearance 3-6 weeks post-surgery is associated with improved EFS and OS in contrast to persistent ctDNA positivity. Pre-surgery and early post-surgery ctDNA measurements independently associated with EFS and OS in exploratory multivariate analyses. Taken together, these data support a prognostic role for serial ctDNA measurement in OAC. Moreover, we show that perioperative ctDNA status adds to the prognostic value of TRG, with a higher risk of death seen in patients whose tumours had a TRG4/5 response to neoadjuvant chemotherapy and who did not clear ctDNA after surgery versus those who do clear ctDNA or who were persistently negative. However, whilst the data presented in this non-randomised observational study suggest that chemotherapy may associate with better outcomes for patients with a positive early post-surgery ctDNA measurement, these data are heavily confounded by small patient numbers and bias in the selection of patients who received adjuvant treatment.

ctDNA positivity at 3-6 months strongly associated with the likelihood of a death or recurrence event occurring within a year. There did not appear to be a relationship between patterns in 3-6 month surveillance ctDNA-positivity and whether recurrence was locoregional or metastatic. However, these data are limited by a reliance on CT for surveillance, given the known poor sensitivity of this modality for identifying locoregional recurrence, as well as by poorly standardised surveillance regimes across recruiting centres. Given that events occurred 10-times earlier in those with a positive 3-6 month ctDNA test versus those who were negative, 71.4% of whom had been ctDNA positive prior to surgery, these data also suggest that serial rather than single timepoint ctDNA assessment is likely to be advantageous in the surveillance setting.

Together, these data add to a growing body of evidence that suggests a role for ctDNA evaluation in prognosticating outcomes in gastroesophageal adenocarcinomas^15–18,20^. In the recent PLAGAST study, blood samples collected from patients with gastric or gastroesophageal junctional cancer before, during and after neoadjuvant therapy, as well as after surgery, were retrospectively analysed with the same personalised tumour-informed ctDNA assay used in this study^15^. In PLAGAST, ctDNA positivity was associated with significantly worse EFS and OS when measured during and after neoadjuvant therapy, and after surgery. Moreover, clearance of ctDNA during neoadjuvant therapy was associated with improved EFS and OS. However, PLAGAST did not directly report on the prognostic value of temporal trends in ctDNA taken immediately prior to, and in the early period following, surgical resection.

Others have similarly demonstrated the prognostic value of pre-operative tumour-informed ctDNA testing. In the phase II PANDA trial, pre-surgical ctDNA concentration correlated with pathological response to neoadjuvant chemoimmunotherapy in patients with gastric and gastroesophageal junctional adenocarcinoma, whilst pre-surgical ctDNA positivity was shown to associate with risk of recurrence^16^. In the context of OAC, our group has previously shown that ctDNA positivity when assessed before surgery was associated with poorer disease-free survival (DFS), albeit in a limited retrospective cohort of 20 patients^17^. However, in this analysis no patients who were ctDNA negative prior to surgery recurred. In the larger prospective cohort reported here, 12.5% (3/24) of patients with ctDNA-negative results at both perioperative ctDNA sampling points recurred, though these cases were marked by other high risk features, including nodal disease at presentation and resection, and a TRG of 3 or 4. Notably, one patient with negative pre-surgery ctDNA turned positive after surgery, whilst another developed a positive ctDNA test 5.8 months after surgery. There were no events in patients with ypT1/2 or ypN0 disease who were persistently ctDNA-negative at both perioperative timepoints.

Use of a tumour-naïve, fixed cancer gene panel to detect ctDNA highlighted an association between pre-operative positivity and the odds of recurrence in patients with gastric cancer enrolled in the phase III CRITICS trial^21^. Similarly, we have used a tumour-naïve panel to demonstrate an association between postoperative ctDNA positivity and shorter DFS and OS in OAC^17,22^. Tumour-informed analysis of early post-operative ctDNA concentration has also demonstrated an association with recurrence risk in a retrospective analysis of patients with OAC, albeit in a heterogenous cohort of around 25 patients^18^. This and other analyses also demonstrated the potential for later ctDNA measurements to identify recurrence sooner than it is recognised by conventional surveillance regimes.

Building on this existing evidence, our study provides prospective data from multiple centres within a single National Health Service that illustrate the prognostic potential of trends in serial perioperative ctDNA measurements in OAC. However, the cohort summarised here included only those patients with microsatellite stable disease and cannot therefore be extrapolated to the approximately 4-5% of patients with microsatellite instability high OAC^23^. TRG data are also prone to inter-observer variability because we did not mandate central pathology review. Moreover, this work was exploratory and the sample size is not based on a formal power calculation. Consequently, this study is underpowered for formal evaluation of the potential for ctDNA testing in this setting and the data should be seen as hypothesis-generating. This includes the included multivariate analyses, which may be unstable and are prone to overfitting. The median follow-up time of 25 months is also relatively short for evaluating OS, which complicates evaluation of the longer-term prognostic value of ctDNA detection.

This analysis additionally provides evidence relating to the feasibility of the commercial assay, with high patient compliance and results returned at a median of 11 days following sample submission where a panel was generated. This timeframe is sufficient for clinical decision making, not least given that adjuvant chemotherapy would generally commence more than two weeks and up to 12 weeks following oesophagectomy. However, this is caveated by a failure to generate bespoke ctDNA assays due to insufficient cellularity for 10% of the initially consented cohort. Moreover, 11% of patients who consented for the study demonstrated a pCR and could not therefore have tumour-informed assays generated from a resection specimen, and as such the cohort is enriched for those who did not have a pCR. Patients for whom a panel could or could not be generated nevertheless otherwise share similar clinicopathological characteristics. Following a protocol amendment, lymph node tissue was evaluated in a small number of patients but did not meet size or cellularity requirements. Routine diagnostic biopsies were similarly too small for analysis. It is not clear what proportion of positive lymph nodes in patients for whom tumour specimens were inadequate could have been successfully used to generate a bespoke ctDNA panel.

Overall, this exploratory prospective multi-centre study provides evidence to support further, powered, studies of a tumour-informed ctDNA assay to guide treatment decisions in patients with OAC. This should include assessments of the ability of ctDNA to guide adjuvant therapy recommendations in the era of perioperative chemoimmunotherapy, as well as an evaluation of the potential for the early identification of recurrence to improve OS, such as by permitting treatment of an oligometastatic state^24^. In particular, the data here point to potential benefit from serial perioperative testing, though the value of this compared with single timepoint assessment has not been formally tested. However, given that around 5-10% of patients who have a pCR subsequently recur^25^, there is a need to evaluate whether nodal or biopsy tissue can be used to generate ctDNA panels in patients for whom a pCR has been achieved.

## Data Availability

Data produced in the present study are available upon reasonable request to the authors but will not be made available for commercial use.

https://github.com/fitzgerald-lab/Periop_ctDNA

## ADDITIONAL INFORMATION

### Data availability

Non-proprietary data are available on reasonable request.

## Acknowledgements

The authors are grateful to Mr Joe Paul (Nottingham University Hospitals NHS Trust) for their support in collating data for this manuscript. Commercial ctDNA assays and related tumour sequencing were provided without charge by Natera. Natera were not supplied with study data in advance of manuscript submission and had no role in data analysis or interpretation. A copy of this manuscript and its subsequent revisions were provided to Natera prior to submission and resubmission. The Oesophageal Cancer Clinical & Molecular Stratification (OCCAMS) study and its successor OCCAMS 2 study were funded by Programme Grant funding (A15874, A22720, A22131) from Cancer Research UK (CRUK). RCF was a recipient of a programme grant from the Medical Research Council (MR/W014122/1) for the duration of this study. CMJ was supported for the duration of this work by a Clinical Lectureship part-funded by CRUK RadNet Cambridge (C17918/A28870), the Loke Medical Sciences Fellowship funded by King’s College, Cambridge, and a CRUK Clinician Scientist Fellowship (RCCCSF-May26/100001). The authors wish to extend their thanks to the patients who enrolled in this study and the clinical teams who supported their participation. The authors are also grateful to the Human Research Tissue Bank at Addenbrooke’s Hospital, Cambridge University Hospitals NHS Foundation Trust, and for the support provided to it by the National Institute for Health & Care Research (NIHR) Cambridge Biomedical Research Centre (BRC-1215-200014). The views expressed are those of the authors and not necessarily those of the NIHR or the Department of Health and Social Care.

## Author contributions

HRC and AF oversaw and supported study setup and study progress, sample processing, study administration, data collection, data cleaning and data analyses. DHJ performed analyses and supported data interpretation. NG, BN and CM supported sample processing and study administration. NG, BN, AH, JHS, JG, RM, LM and SLP consented participants, undertook collection of clinicopathological, treatment and outcomes data, and contributed to interpretation of the study results. GD, VA and SM supported data and sample processing. RCF conceived, obtained funding for, and oversaw the study. CMJ supported collection of clinicopathological, treatment and outcomes data, undertook study oversight, contributed to data cleaning, led study analyses, supported data interpretation and wrote the first draft of the manuscript. All authors have read and contributed to revisions of the manuscript, and all agree with the final version.

---

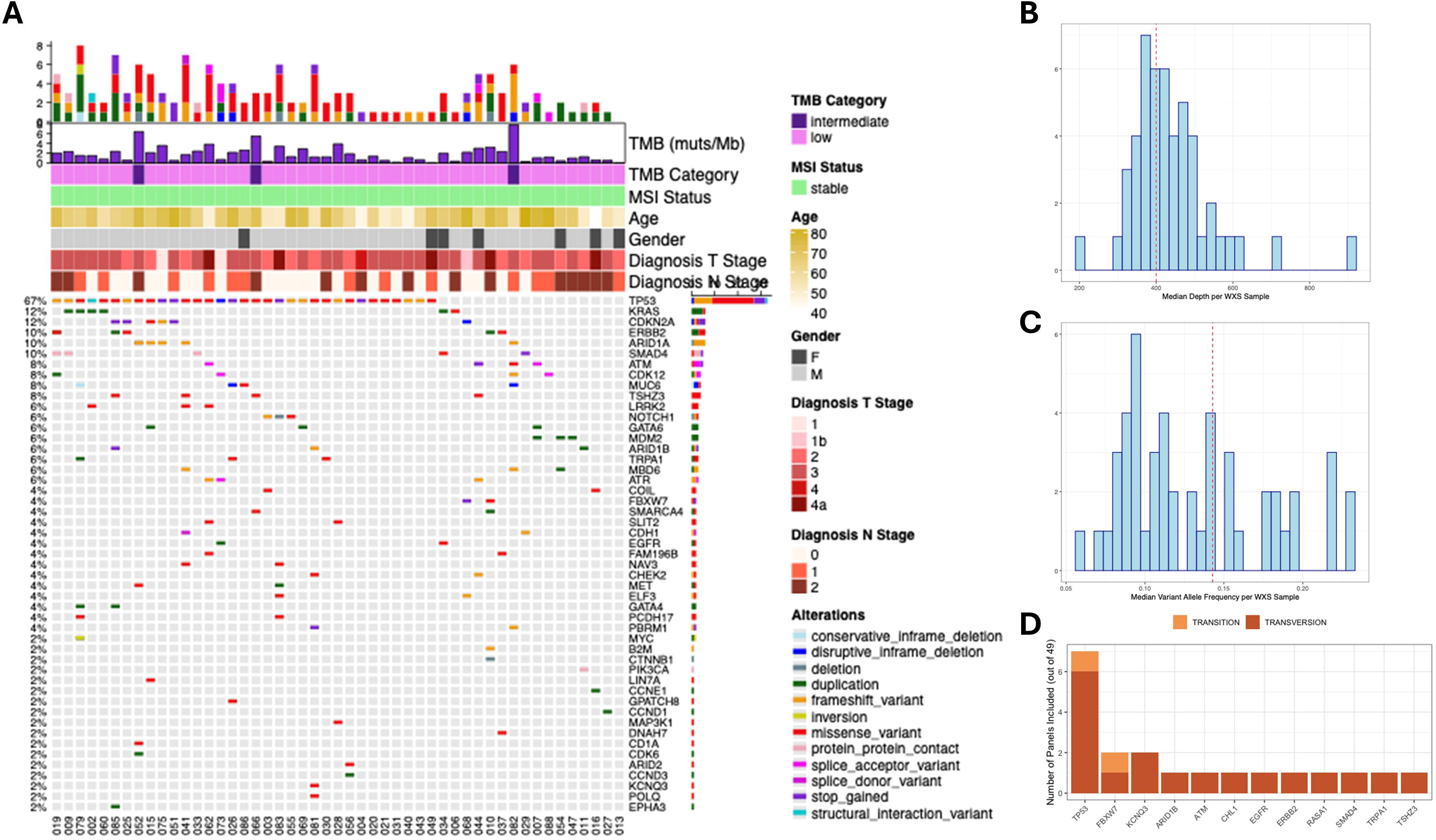

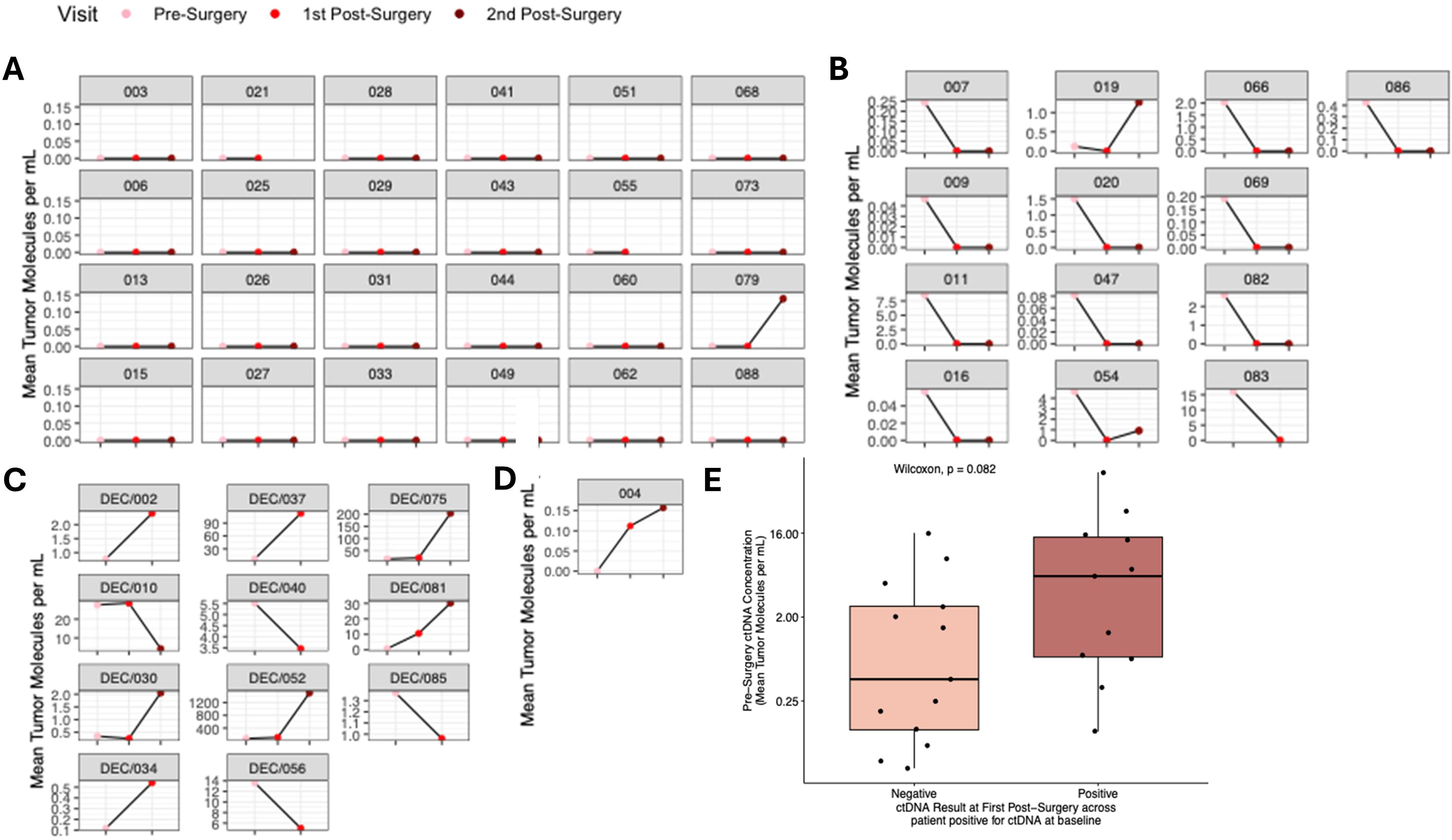

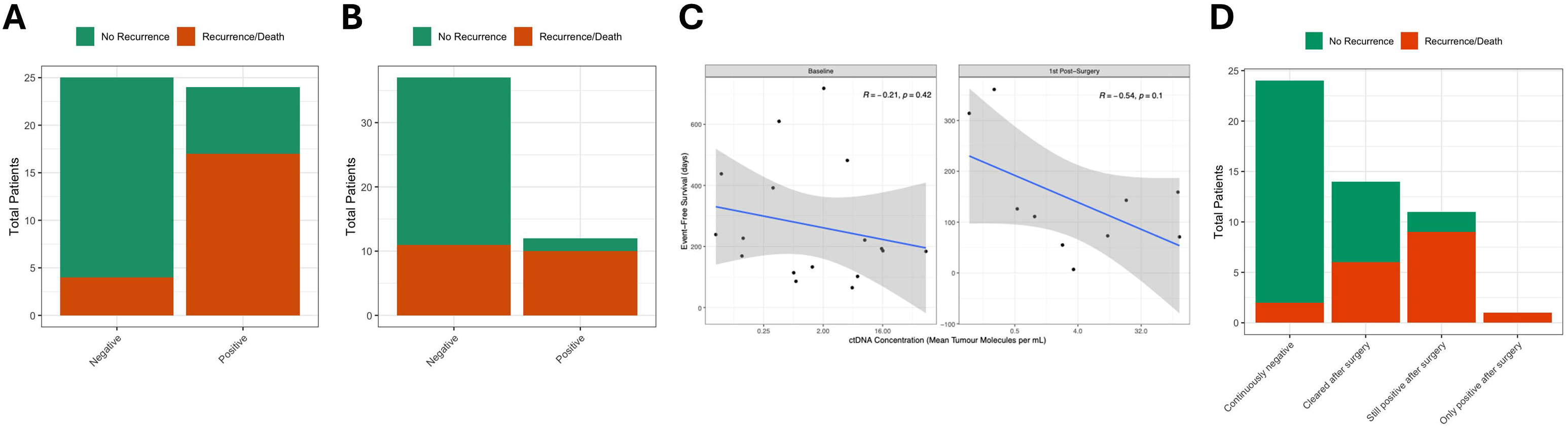

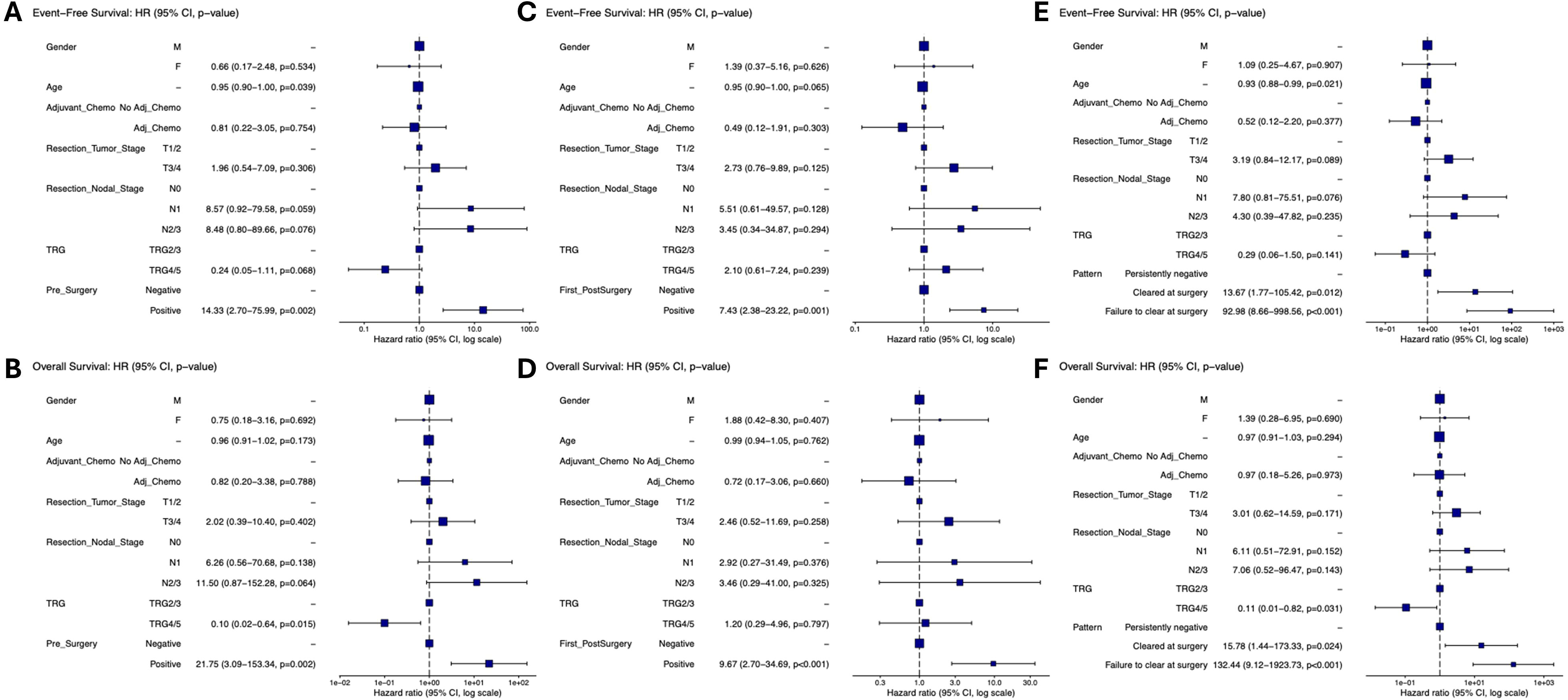

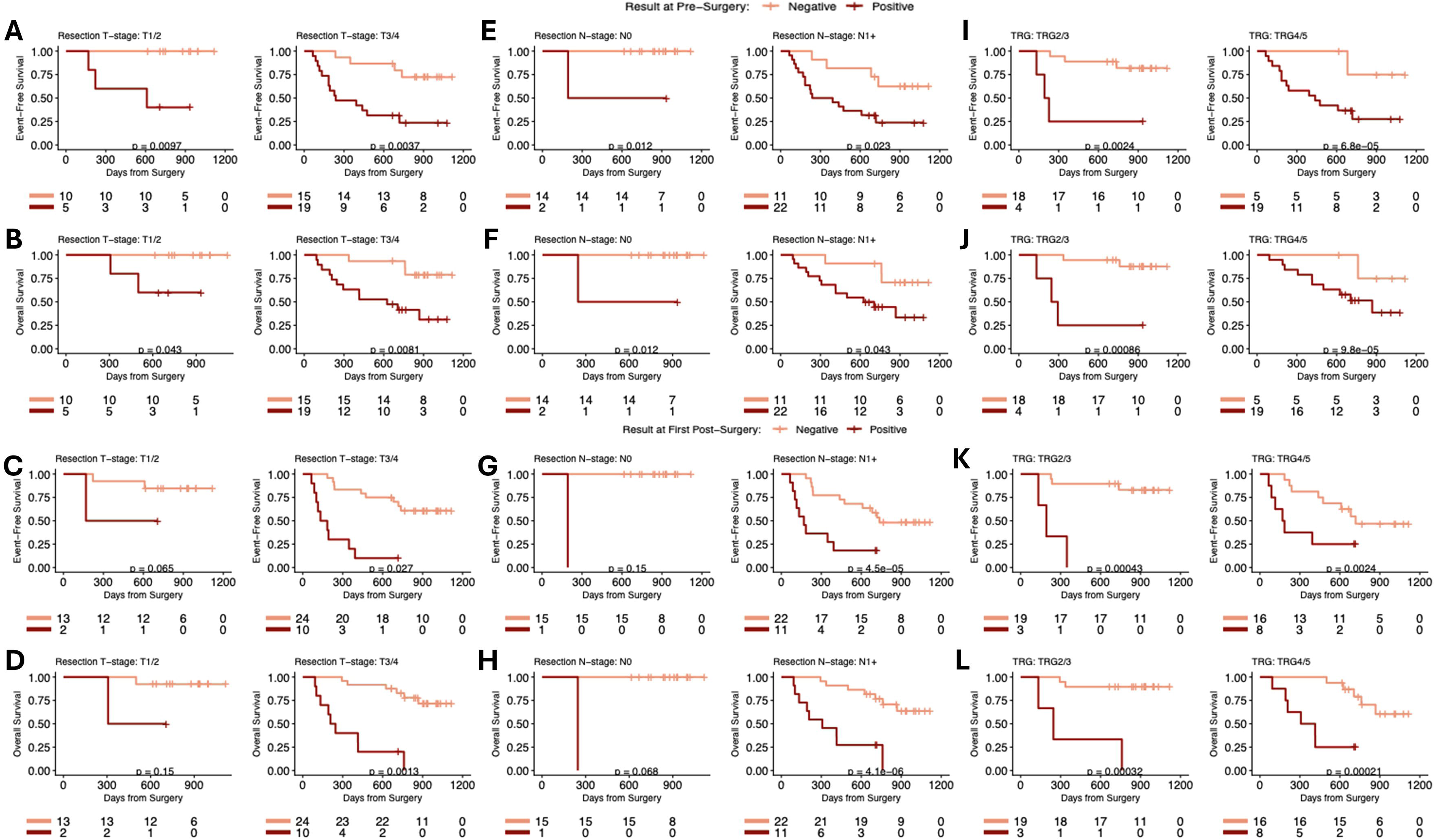

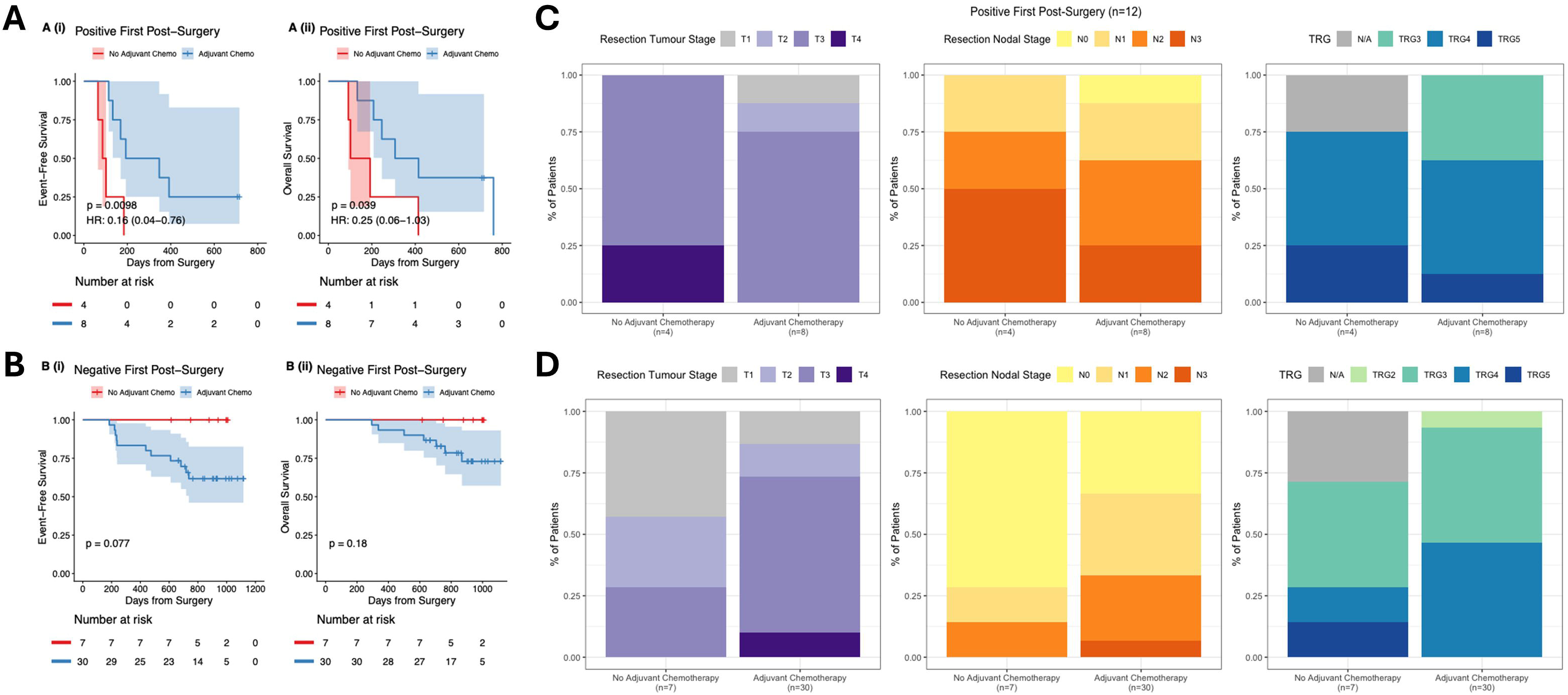

## REFERENCES

1. Morgan E, Soerjomataram I, Rumgay H, et al. The Global Landscape of Esophageal Squamous Cell Carcinoma and Esophageal Adenocarcinoma Incidence and Mortality in 2020 and Projections to 2040: New Estimates From GLOBOCAN 2020. Gastroenterology 2022; 163(3): 649–58.e2.

2. Then EO, Lopez M, Saleem S, et al. Esophageal Cancer: An Updated Surveillance Epidemiology and End Results Database Analysis. World J Oncol 2020; 11(2): 55–64.

3. Hoeppner J, Brunner T, Schmoor C, et al. Perioperative Chemotherapy or Preoperative Chemoradiotherapy in Esophageal Cancer. N Engl J Med 2025; 392(4): 323–35.

4. Cunningham D, Allum WH, Stenning SP, et al. Perioperative chemotherapy versus surgery alone for resectable gastroesophageal cancer. N Engl J Med 2006; 355(1): 11–20.

5. Janjigian YY, Al-Batran SE, Wainberg ZA, et al. Perioperative Durvalumab in Gastric and Gastroesophageal Junction Cancer. N Engl J Med 2025; 393(3): 217–230.

6. Lordick F, Mauer ME, Stocker G, et al. Adjuvant immunotherapy in patients with resected gastric and oesophagogastric junction cancer following preoperative chemotherapy with high risk for recurrence (ypN+ and/or R1): European Organisation of Research and Treatment of Cancer (EORTC) 1707 VESTIGE study. Ann Oncol 2025; 36(2): 197–207.

7. SPACE-FLOT Investigators. Pathological response guides adjuvant 5-fluorouracil, leucovorin, oxaliplatin, and docetaxel (FLOT) chemotherapy in surgically resected gastro-oesophageal cancer (SPACE-FLOT): international cohort study. Br J Surg 2025; 112(4):znaf056.

8. Mandard AM, Dalibard F, Mandard JC, et al. Pathologic assessment of tumor regression after preoperative chemoradiotherapy of esophageal carcinoma. Clinicopathologic correlations. Cancer 1994; 73(11): 2680–6.

9. Knight WRC, Zylstra J, Van Hemelrijck M, et al. Patterns of recurrence in oesophageal cancer following oesophagectomy in the era of neoadjuvant chemotherapy. BJS Open 2017; 1(6): 182–90.

10. Peyre CG, Hagen JA, DeMeester SR, et al. Predicting systemic disease in patients with esophageal cancer after esophagectomy: a multinational study on the significance of the number of involved lymph nodes. Ann Surg 2008; 248(6): 979–85.

11. Lagarde SM, Phillips AW, Navidi M, Disep B, Immanuel A, Griffin SM. The presence of lymphovascular and perineural infiltration after neoadjuvant therapy and oesophagectomy identifies patients at high risk for recurrence. Br J Cancer 2015; 113(10): 1427–33.

12. von Rahden BH, Stein HJ, Feith M, Becker K, Siewert JR. Lymphatic vessel invasion as a prognostic factor in patients with primary resected adenocarcinomas of the esophagogastric junction. J Clin Oncol 2005; 23(4): 874–9.

13. Jayaprakasam VS. Yeh R, Ku GY, Petkovska I, Fuqua JL Gollub M, Paroder V. Role of imaging in esophageal cancer management in 2020: update for radiologists. AJR Am J Roentgenol 2020;215(5):1072–1084.

14. Lockwood CM, Messersmith HJ, Kim AS, et al. Circulating Tumor DNA Testing in Solid Tumors and Lymphoma: ASCO Guideline. JCO Oncol Pract 2026: 22(9):1468–1489.

15. Zaanan A, Didelot A, Broudin C, et al. Longitudinal circulating tumor DNA analysis during treatment of locally advanced resectable gastric or gastroesophageal junction adenocarcinoma: the PLAGAST prospective biomarker study. Nat Commun 2025; 16(1): 6815.

16. Verschoor YL, van de Haar J, van den Berg JG, et al. Neoadjuvant atezolizumab plus chemotherapy in gastric and gastroesophageal junction adenocarcinoma: the phase 2 PANDA trial. Nat Med 2024; 30(2): 519–30.

17. Ococks E, Sharma S, Ng AWT, Aleshin A, Fitzgerald RC, Smyth E. Serial Circulating Tumor DNA Detection Using a Personalized, Tumor-Informed Assay in Esophageal Adenocarcinoma Patients Following Resection. Gastroenterology 2021; 161(5): 1705–8.e2.

18. Huffman BM, Aushev VN, Budde GL, et al. Analysis of Circulating Tumor DNA to Predict Risk of Recurrence in Patients With Esophageal and Gastric Cancers. JCO Precis Oncol 2022; 6: e2200420.

19. Reinert T, Henriksen TV, Christensen E, et al. Analysis of Plasma Cell-Free DNA by Ultradeep Sequencing in Patients With Stages I to III Colorectal Cancer. JAMA Oncol 2019; 5(8): 1124–31.

20. Mehta R, Rivero-Hinojosa S, Dayyani F, et al. Circulating tumor DNA informs clinical practice in patients with recurrent/metastatic gastroesophageal cancers. Cancer 2026; 132(1): e70242.

21. Leal A, van Grieken NCT, Palsgrove DN, et al. White blood cell and cell-free DNA analyses for detection of residual disease in gastric cancer. Nat Commun 2020; 11(1): 525.

22. Ococks E, Frankell AM, Masque Soler N, et al. Longitudinal tracking of 97 esophageal adenocarcinomas using liquid biopsy sampling. Ann Oncol 2021; 32(4): 522–32.

23. Okui J, Prakash LR, Lyu GH et al. Immunotherapy response in microsatellite instability-high gastroesophageal adenocarcinoma. JAMA Surg 2026; 161(6): 600–7.

24. Cammarota A, Puccini A, Jones CM, van Rossum PSN, Mouliere F, van Laarhoven HWM. Circulating tumour DNA in oligometastatic oesophago-gastric cancers: applications, challenges, and future directions. Dis Esophagus 2026; 39(3).

25. Alhayo S, Chou WK, Giorgi L et al. FLOT vs CROSS among patients with pathological complete response in esophageal adenocarcinoma. JAMA Surg 2026; 161(9): 917–925.

