## Supplementary file for "Molecular residual disease detection by serial tumour-informed circulating tumour DNA analysis in resectable oesophageal & gastroesophageal junctional adenocarcinoma: a prospective UK multi-centre study"

**SUPPLEMENTARY TABLES**

| **Site** | **Trust** | **Principal investigator** |
| --- | --- | --- |
| Addenbrooke’s Hospital | Cambridge University Hospitals NHS Foundation Trust | Prof. Rebecca Fitzgerald |
| Salford Royal Hospital | Northern Care Alliance NHS Foundation Trust | Mr John Saunders |
| Nottingham University Hospital | Nottingham University Hospitals NHS Trust | Mr Simon Parsons |
| St Thomas Hospital | Guy’s and St Thomas’ NHS Foundation Trust | Mr James Gossage |

**Supplementary table 1: A summary of participating English National Health Service centres.**

| **Reason for exclusion** | | **No.** |
| --- | --- | --- |
| Inability to perform whole exome sequencing on the resection specimen | | |
|  | Complete pathological response | 9 |
|  | Insufficient cellularity | 10 |
|  | Failed germline extraction | 1 |
| Failure to proceed to, or complete, curative resection | |  |
|  | Disease progression/unresectable | 5 |
|  | Surgical complication | 1 |
|  | Death following surgery | 1 |
| Change in tumour classification | |  |
|  | Histological type changed to neuroendocrine based on analysis of resection specimen | 1 |
| Logistical issues | |  |
|  | Did not attend for repeat sampling or sample could not be collected | 6 |
|  | Initial ctDNA measurement taken outside of the perioperative timeframe (i.e. surgery delayed) | 1 |
|  | Failed sample shipment | 2 |
| Withdrawal of, or inability to provide, consent | |  |
|  | Perioperative morbidity (prolonged ITU stay) with no capacity to consent for sampling | 1 |
|  | Active withdrawal of consent | 2 |

**Supplementary table 2: a list of reasons for exclusion from the final study analysis.**

|  | | | | **n=36*** | **%** |
| --- | --- | --- | --- | --- | --- |
| **Patient characteristics** | | | |  |  |
|  | Age (4 unknown) | | |  |  |
|  |  | 40-49 | | 3 | 8.3 |
|  |  | 50-59 | | 8 | 22.2 |
|  |  | 60-69 | | 11 | 30.6 |
|  |  | 70-79 | | 9 | 25.0 |
|  |  | 80-89 | | 1 | 2.8 |
|  | Sex | | |  |  |
|  |  | Male | | 29 | 80.6 |
|  |  | Female | | 7 | 19.4 |
|  | ECOG performance status (3 unknown) | | |  |  |
|  |  | 0 | | 22 | 61.1 |
|  |  | 1 | | 11 | 30.6 |
| **Tumour characteristics** | | | |  |  |
|  | Tumour location (7 unknown) | | |  |  |
|  |  | Middle oesophageal | | 1 | 2.8 |
|  |  | Lower oesophageal | | 8 | 22.2 |
|  |  | GOJ | |  |  |
|  |  |  | Unclassified | 2 | 5.6 |
|  |  |  | Siewert I | 8 | 22.2 |
|  |  |  | Siewert II | 7 | 19.4 |
|  |  |  | Siewert III | 1 | 2.8 |
|  | Tumour differentiation** (19 unknown) | | |  |  |
|  |  | Well | | 1 | 2.8 |
|  |  | Moderate | | 7 | 19.4 |
|  |  | Poor | | 9 | 25 |
|  | Clinical T-stage (8 unknown) | | |  |  |
|  |  | 0 | | 7 | 19.4 |
|  |  | 1 | | 7 | 19.4 |
|  |  | 2 | | 4 | 11.1 |
|  |  | 3 | | 10 | 27.8 |
|  | Clinical N-stage (8 unknown) | | |  |  |
|  |  | 0 | | 18 | 50.0 |
|  |  | 1 | | 6 | 16.7 |
|  |  | 2 | | 3 | 8.3 |
| **Outcomes** | | | | | |
|  | TRG*** | | |  |  |
|  |  | 1 | | 10 | 27.8 |
|  |  | 2 | | 2 | 5.3 |
|  |  | 3 | | 10 | 27.8 |
|  |  | 4 | | 7 | 18.4 |
|  |  | 5 | | 0 | 0.0 |

**Supplementary table 3: clinicopathological and treatment-related outcomes for patients for whom ctDNA panels could not be generated.** *Three patients who withdrew consent to study participation, and one patient for whom diagnosis was revised to an oesophageal neuroendocrine tumour, are not included in these data. **Most severe classification shown. ***Seven patients did not proceed to resection despite receipt of neoadjuvant therapy.

| **Treatment characteristics** | | | | **n=49** | **%** |
| --- | --- | --- | --- | --- | --- |
|  | Pre-operative FLOT – cycles: | | |  |  |
|  |  | 1 | | 1 | 2.0 |
|  |  | 3 | | 1 | 2.0 |
|  |  | 4 | | 44 | 91.8 |
|  |  | None | | 3 | 6.1 |
|  | Post-operative FLOT – cycles: | | |  |  |
|  |  | 1 | | 5 | 10.2 |
|  |  | 2 | | 1 | 2.0 |
|  |  | 3 | | 6 | 12.2 |
|  |  | 4 | | 26 | 53.1 |
|  |  | None | | 11 | 22.5 |
| **Outcomes** | | | | **n=46*** | **%** |
|  | Mandard TRG | | |  |  |
|  |  | 2 | | 2 | 4.3 |
|  |  | 3 | | 20 | 43.5 |
|  |  | 4 | | 21 | 45.7 |
|  |  | 5 |  | 3 | 6.5 |

**Supplementary table 4: treatment characteristics and pathological outcomes.** FLOT: 5-fluorouracil, leucovorin, oxaliplatin, docetaxel; TRG: tumour regression grade. *Three of the 49 patients enrolled in the full cohort did not receive neoadjuvant therapy.

|  | **Pre-surgery** | | | | **Early post-surgery** | | | |
| --- | --- | --- | --- | --- | --- | --- | --- | --- |
|  | **Negative**  n = 25*^1^* | **Positive**  n = 24*^1^* | **p-value***^2^* | **q-value***^3^* | **Negative**  n = 37*^1^* | **Positive**  n = 12*^1^* | **p-value***^4^* | **q-value***^3^* |
| **Gender** |  |  | 0.7 | 0.8 |  |  | 0.7 | 0.8 |
| Female | 3 (12%) | 4 (17%) |  |  | 6 (16%) | 1 (8.3%) |  |  |
| Male | 22 (88%) | 20 (83%) |  |  | 31 (84%) | 11 (92%) |  |  |
| **Performance status pre-treatment*** |  |  | 0.5 | 0.8 |  |  | 0.7 | 0.8 |
| 0 | 15 (63%) | 17 (71%) |  |  | 23 (64%) | 9 (75%) |  |  |
| 1 | 9 (38%) | 7 (29%) |  |  | 13 (36%) | 3 (25%) |  |  |
| **Anatomical location** |  |  | 0.077 | 0.2 |  |  | 0.2 | 0.4 |
| GOJ | 4 (16%) | 2 (8.3%) |  |  | 5 (14%) | 1 (8.3%) |  |  |
| Lower Oesophagus | 4 (16%) | 5 (21%) |  |  | 6 (16%) | 3 (25%) |  |  |
| Siewert 1 | 8 (32%) | 2 (8.3%) |  |  | 10 (27%) | 0 (0%) |  |  |
| Siewert 2 | 9 (36%) | 11 (46%) |  |  | 13 (35%) | 7 (58%) |  |  |
| Siewert 3 | 0 (0%) | 4 (17%) |  |  | 3 (8.1%) | 1 (8.3%) |  |  |
| **Differentiation status**** |  |  | 0.9 | 0.9 |  |  | 0.2 | 0.4 |
| Moderate | 10 (42%) | 9 (39%) |  |  | 17 (47%) | 2 (18%) |  |  |
| Poor | 14 (58%) | 14 (61%) |  |  | 19 (53%) | 9 (82%) |  |  |
| **Resection T stage** |  |  | 0.12 | 0.2 |  |  | 0.8 | 0.8 |
| T1 | 7 (28%) | 1 (4.2%) |  |  | 7 (19%) | 1 (8.3%) |  |  |
| T2 | 3 (12%) | 4 (17%) |  |  | 6 (16%) | 1 (8.3%) |  |  |
| T3 | 14 (56%) | 16 (67%) |  |  | 21 (57%) | 9 (75%) |  |  |
| T4 | 1 (4.0%) | 3 (13%) |  |  | 3 (8.1%) | 1 (8.3%) |  |  |
| **Resection N stage** |  |  | <0.001 | 0.002 |  |  | 0.031 | 0.2 |
| N0 | 14 (56%) | 2 (8.3%) |  |  | 15 (41%) | 1 (8.3%) |  |  |
| N1 | 6 (24%) | 8 (33%) |  |  | 11 (30%) | 3 (25%) |  |  |
| N2 | 5 (20%) | 8 (33%) |  |  | 9 (24%) | 4 (33%) |  |  |
| N3 | 0 (0%) | 6 (25%) |  |  | 2 (5.4%) | 4 (33%) |  |  |
| **TRG** |  |  | <0.001 | <0.001 |  |  | 0.2 | 0.4 |
| TRG2 | 2 (8.7%) | 0 (0%) |  |  | 2 (5.7%) | 0 (0%) |  |  |
| TRG3 | 16 (70%) | 4 (17%) |  |  | 17 (49%) | 3 (27%) |  |  |
| TRG4 | 5 (22%) | 16 (70%) |  |  | 15 (43%) | 6 (55%) |  |  |
| TRG5 | 0 (0%) | 3 (13%) |  |  | 1 (2.9%) | 2 (18%) |  |  |

**Supplementary table 5: Association between pre-surgery and early post-surgery ctDNA levels, and clinicopathological characteristics.**. ^1^n (%). ^2^Fisher's exact test; Pearson's Chi-squared test. ^3^False discovery rate correction for multiple testing. *Unknown in one patient. **Unknown for two patients. ECOG: Eastern Cooperative Oncology Group Performance Status; N-stage: Nodal stage; T-stage: tumour stage; TRG: (Mandard) tumour regression grade.

|  |  |  |  | **Event-free survival** | | **Overall survival** | |
| --- | --- | --- | --- | --- | --- | --- | --- |
|  |  |  | **Total (Events, Deaths)** | **HR** | **p-value** | **HR** | **p-value** |
| **ypT-stage** | |  |  |  |  |  |  |
|  | T1/2 | Cleared at surgery | 3 (2, 1) |  |  |  |  |
|  | T1/2 | Persistently negative | 10 (0, 0) | - | - | - | - |
|  |  | Failure to clear | 2 (1, 1) | 1.03 | 0.984 | 2.12 | 0.600 |
|  | T3/4 | Cleared at surgery | 10 (6, 4) |  |  |  |  |
|  | T3/4 | Persistently negative | 14 (3, 2) | 0.25 | 0.052 | 0.30 | 0.162 |
|  |  | Failure to clear | 9 (8, 8) | 4.63 | 0.008 | 8.15 | 0.002 |
| **ypN-stage** | |  |  |  |  |  |  |
|  | N0 | Cleared at surgery | 1 (0, 0) |  |  |  |  |
|  | N0 | Persistently negative | 14 (0, 0) | - | - | - | - |
|  |  | Failure to clear | 1 (1, 1) | - | - | - | - |
|  | N1+ | Cleared at surgery | 12 (8, 5) |  |  |  |  |
|  | N1+ | Persistently negative | 10 (3, 2) | 0.31 | 0.088 | 0.38 | 0.250 |
|  |  | Failure to clear | 10 (8, 8) | 2.91 | 0.041 | 5.36 | 0.007 |
| **TRG** | |  |  |  |  |  |  |
|  | TRG2/3 | Cleared at surgery | 2 (1, 1) |  |  |  |  |
|  | TRG2/3 | Persistently negative | 17 (2, 1) | 0.16 | 0.144 | 0.08 | 0.083 |
|  |  | Failure to clear | 2 (2, 2) | - | - | - | - |
|  | TRG4/5 | Cleared at surgery | 11 (7, 4) |  |  |  |  |
|  | TRG4/5 | Persistently negative | 5 (1, 1) | 0.24 | 0.177 | 0.43 | 0.456 |
|  |  | Failure to clear | 8 (6, 6) | 2.44 | 0.122 | 6.79 | 0.010 |

**Supplementary table 6: Prognostic value of perioperative ctDNA patterns for predicting event free and overall survival.** All hazard ratios (HRs) calculated via univariate analysis in comparison to the ‘Cleared at surgery’ group, which was used as a reference. Missing values (“-”) indicate tests on data with complete separation arising due to insufficient events. TRG: tumour regression grade.

**LIST OF SUPPLEMENTARY FIGURES**

**Supplementary figure 1: Genomic features of the studied cohort.** **(A)** Oncoplot demonstrating the relationship between the mutational profile for each sequenced tumour and the clinicopathological characteristics of the related patient. **(B)** Frequency histogram demonstrating median sequencing depths achieved for each sample. **(C)** Frequency histogram depicting the median variant allele frequency per sample. **(D)** Frequencies of inclusion of OAC-related driver genes in patient-specific panels. MSI: microsatellite instability; TMB: tumour mutational burden.

**Supplementary figure 2: Per-patient trends in ctDNA positivity at each of the studied timepoints & their relationship to pre-surgery ctDNA concentration. (A)** Continuously negative across all three timepoints (n=24). **(B)** Clearance at surgery (n=13). **(C)** Persistent positivity pre- and post-surgery (n=11). **(D)** Newly positive following surgery (n=1). **(E)** A comparison of pre-surgery ctDNA concentration between patients who were negative and positive for ctDNA at the early post-surgery timepoint. Comparison drawn using the Wilcoxon test.

**Supplementary figure 3: Association between ctDNA measurement and event free survival. (A)** Proportion of patients with a negative or positive pre-surgery ctDNA test who subsequently developed recurrence or died. **(B)** Proportion of patients with a negative or positive early post-surgery ctDNA test whos subsequently developed recurrence or died. **(C)** Association between mean ctDNA concentration at pre-surgery or early post-surgery timepoints and event free survival. **(D)** Incidence of events (recurrence or death) by perioperative ctDNA measurement. One patient in the ‘Still positive after surgery’ group died with no known recurrence.

**Supplementary figure 4: Multivariate analyses for event free and overall survival by perioperative ctDNA positivity. (A)** Event free and **(B)** overall survival by pre-surgery ctDNA status. **(C)** Event free and **(D)** overall survival by early post-surgery ctDNA status. **(E)** Event free and **(F)** overall survival by combined perioperative ctDNA status.

**Supplementary figure 5: Event-free and overall survival outcomes by pre- or early post- surgery ctDNA status and pathological indices**. **(A)** Event free and **(B)** overall survival by T-stage and pre-surgery ctDNA status. **(C)** Event free and **(D)** overall survival by T-stage and early post-surgery ctDNA status. **(E)** Event free and **(F)** overall survival by N-stage and pre-surgery ctDNA status. **(G)** Event free and **(H)** overall survival by N-stage and early post-surgery ctDNA status. **(I)** Event free and **(J)** overall survival by TRG and pre-surgery ctDNA status. **(K)** Event free and **(L)** overall survival by TRG and early post-surgery ctDNA status. P-values underwent FDR-correction for multiple testing across event-free and overall survival analyses separately.

**Supplementary figure 6: Event free and overall survival by ctDNA status and receipt of adjuvant chemotherapy. (A)** **(i)** Event free and **(ii)** overall survival by receipt of chemotherapy for patients with a positive early post-surgery ctDNA test. **(B)** **(i)** Event free and **(ii)** overall survival by receipt of chemotherapy for patients with a negative early post-surgery ctDNA test. **(C)** A comparison of the pathological characteristics of patients positive for ctDNA at the early post-surgery timepoint and who received versus did not receive adjuvant chemotherapy. **(D)** A comparison of the pathological characteristics of patients negative for ctDNA at the early post-surgery timepoint and who received versus did not receive adjuvant chemotherapy.
